# Clinical pattern and diagnostic of comorbidity of respiratory tuberculosis and viral pneumonia caused by Herpesvirus Simplex, Human Cytomegalovirus and SARS-CoV-2 in patients with late-stage HIV infection with immunodeficiency

**DOI:** 10.1101/2023.08.30.23294716

**Authors:** V. Yu. Mishin, A.V. Mishina, I. V. Shashenkov

## Abstract

**Aim:** To study features of social status, clinical pattern and diagnosis in cases of comorbidity of respiratory tuberculosis and viral pneumonia caused by *Herpesvirus Simplex* of type 1, *Human Cytomegalovirus* and *SARS-CoV-2* in patients with late-stage HIV infection with immunodeficiency.

**Materials and methods:** The prospective study included 25 patients with comorbid condition of respiratory tuberculosis with *Mycobacterium tuberculosis* in excreta, herpesvirus and coronavirus pneumonia, and 21 patients with respiratory tuberculosis as well as cytomegalovirus and coronavirus pneumonia (1a and 2a main groups) and, respectively, 25 and 21 similar patients, but without coronavirus pneumonia (1b and 2b comparison group) in the late stages of HIV infection with immunodeficiency. For the etiological diagnosis of herpesvirus- and cytomegalovirus pneumonia, the PCR test was used for recognition of DNA of *Herpesvirus Simplex* of type 1 and *Human Cytomegalovirus* in the diagnostic material of respiratory tract and for the etiological diagnosis of coronavirus pneumonia, the PCR for recognition of RNA was used to reveal *SARS–CoV-2*. Statistical analysis of the data was performed by the use of the Microsoft Office Excel 2019 software for calculation of group mean, standard error of mean and confidence interval.

**Results:** The comorbidity of respiratory tuberculosis, herpes-, cytomegalo- and coronavirus pneumonia in patients with late-stage HIV infection in the phase of progression and in the absence of ART was characterized by severe immunodeficiency and generalization of tuberculosis with multiple extrapulmonary lesions. The results displayed similarity of clinical manifestations and visualization of changes in CT-picture in cases of comorbidity the diseases which hampers their recognition due to simultaneous combination of several pathologies with similar clinical manifestations that requires a complex etiological diagnosis of the specific diseases to prescribe a timely comprehensive treatment and reduce lethality in this severe contingent of patients.

**Conclusion:** Patients with respiratory tuberculosis and HIV infection registered in the office of tuberculosis care for HIV-infected individuals in the antituberculosis dispensary represent a group of high risk from COVID-19 infection and CVP disease, and, in cases of combination with severe immunodeficiency, HVP and CMVP, the patients should be regularly subjected to preventive studies for timely detection of COVID-19 for the purpose of their emergency isolation and treatment.

## Rationale

Infectious lung damage of various etiologies is one of the main causes of pneumonia in patients with HIV infection especially in the late stages with immunodeficiency [1-5]. An important role in this category of patients plays tuberculosis if combined with other opportunistic lung infections [6] among which important is a viral pneumonia (VP) etiologically associated with *Herpesvirus Simplex* of type 1 (HVS-1) and *Human Cytomegalovirus* (CMVH) that significantly affects the severity of clinical manifestation and the therapy efficacy because of absence of specific treatment [7-12]

COVID-19 has created certain problems in HIV-infected patients due to severe damage to the respiratory system and development of coronavirus pneumonia (CVP) which is one of the main reasons for hospitalization of HIV-infected patients and the most common cause of their death [13-17]. However, the greatest problems arise in cases of comorbidity of respiratory tuberculosis and herpesvirus pneumonia (HVP) caused by *HVS-1*, cytomegalovirus pneumonia (CMVP) caused by *CMVH* and CVP caused by *SARS-CoV-2* in patients with late-stage HIV infection with immunodeficiency in terms of clinical and radiological manifestations and timely diagnosis for selection of adequate etiological treatment and anti-epidemic measures. The management of this category of patients requires new knowledge on the peculiarities of clinical and radiological manifestations and diagnostics as well as measures of individual and collective protection while there are no such publications in the foreign and national literature.

**The aim of the study** was to study the features of social status, clinic and diagnosis of comorbidity of tuberculosis of the respiratory system (TRS) and viral pneumonia (VP) caused by *HVS–1, CMVH* and *SARS-CoV-2* in patients with late stages of HIV infection with immunodeficiency.

## Materials and methods

The work was carried out at the Department of Phthisiology and Pulmonology of the Federal State Budgetary Educational Institution of Higher Education (FSBEI HE) A.I. Evdokimov Moscow State Medical University of the Ministry of Health of the Russian Federation and at the clinical base of Department of the State Budgetary Healthcare Institution (SBHI) “Professor G.A. Zakharin Tuberculosis Clinical Hospital No. 3” of the Moscow Department of Health.

A prospective study performed included 92 of newly diagnosed patients having tuberculosis of respiratory system (TRS) with *Mycobacterium tuberculosis* (MBT) in excreta, stage 4B of HIV infection in the phase of progression and in the absence of antiretroviral therapy (ART) aged 26-52 years. There were 59 men (64.1±5.0%), 33 women (35.9±5.0%). The patients were divided into 1a and 2a (main) and 1b and 2b (control) groups.

The main group 1a was formed of 25 patients who were diagnosed with comorbidity of TRS, HVP and CVP while main group 2a consisted of 21 patients with diagnosed comorbidity of TRS, CMVP and CVP

The comparison groups 1b and 2b were, respectively, composed of: 25 and 21 patients who were not diagnosed with CVP, they were selected according to the “copy-pair” principle in relation to patients from the main groups 1a and 2a who were almost identical in social, age, sexual, clinical parameters, the stage of HIV infection, the severity of immunodeficiency and absence of other opportunistic lung infections (OIL).

For the etiological diagnosis of TRS, the diagnostic material of the respiratory tract (sputum, bronchoalveolar lavage, biopsy material obtained during bronchoscopy and punctures of intra-thoracic lymph nodes) and other organs (blood, urine, feces and punctures of peripheral lymph nodes) was seeded on a dense Lowenstein-Jensen medium and in the automated BACTEC MGIT 960 system with determination of drug resistance of the resulting MBT culture to anti-tuberculosis drugs by the method of absolute concentrations [18].

For the etiological diagnosis of HVP and CMVP, the PCR test for recognition of DNA of *HVS-1* and *CMVH* as well as the ELISA test for detection of viral antigens and antibodies in the diagnostic material of the respiratory tract were used [19], while for the etiological diagnosis of CVP from patients with COVID-19, the PCR for detection of *SARS-CoV-2* RNA was employed [20].

To exclude other OIL pathogens most often developing in the late stages of HIV infection, such as bacterial pneumonia caused by *Streptococcus pneumoniae, Haemophilus influenzae*, or *Staphylococcus aureus (S. aureus), Mycobacterium nontuberculosis, Pneumocystis pneumonia - Pneumocystis jirovecii* or candidiasis pneumonia – *Candida albicans*, the microbiological, immunological methods and the PCR test of diagnostic material from the respiratory tract [19] were used.

All patients underwent complex clinical, laboratory, immunological (assessment of CD4+ lymphocytes number by flow cytofluorometry and viral load by the number of HIV RNA copies in peripheral blood) and radiation examination, which included computed tomography (CT), magnetic resonance imaging (MRI) and ultrasound imaging (UI) of the chest and internal organs.

Statistical analysis of the data was performed by the use of the Microsoft Office Excel 2019 software for calculation of group mean, standard error of mean and confidence interval (CI). The P criterion for statistically significant difference was determined by the Student’s table. The differences between means were considered statistically significant at a value of p<0.05.

## Results

In all 92 patients of the groups selected for analysis, the HIV infection was the first disease, and TRS was detected 5-8 years later. All of them were registered at the AIDS center which they practically did not visit due to social maladaptation and lack of commitment to examination and treatment, they were unemployed and did not have a family. All patients suffered from drug addiction, consumed alcoholic beverages and smoked tobacco products. All patients were diagnosed with concomitant pathology: viral hepatitis B or C and chronic obstructive pulmonary disease (COPD) – in 61 (66.3±4.9%).

TRS was diagnosed in all 92 patients when they applied with symptoms of acute inflammatory respiratory disease in a primary health care institutions and the diagnosis was confirmed by a complex examination in an antituberculosis dispensary (ATD) where a microbiological examination of the diagnostic material from respiratory tract had recognized the MBT and the CT examination of chest had revealed bilateral changes typical for inflammatory lung lesions, so they were hospitalized in the Professor G.A. Zakharin Tuberculosis Clinical Hospital No. 3. In the reception department, 46 patients of groups 1a and 2a were diagnosed with COVID-19 and they were isolated in the observation department (“red zone”) while if COVID-19 not revealed the 46 patients of groups 1b and 2b were referred to the department for patients with tuberculosis and HIV infection where additionally in 50 patients of groups 1a and 1b the *HVS-1* was also detected and in 42 – the *CMVH*.

The distribution of patients in groups of the study by number of CD4+ lymphocytes per 1 uL of blood is shown in Table 1.

**Table 1.**
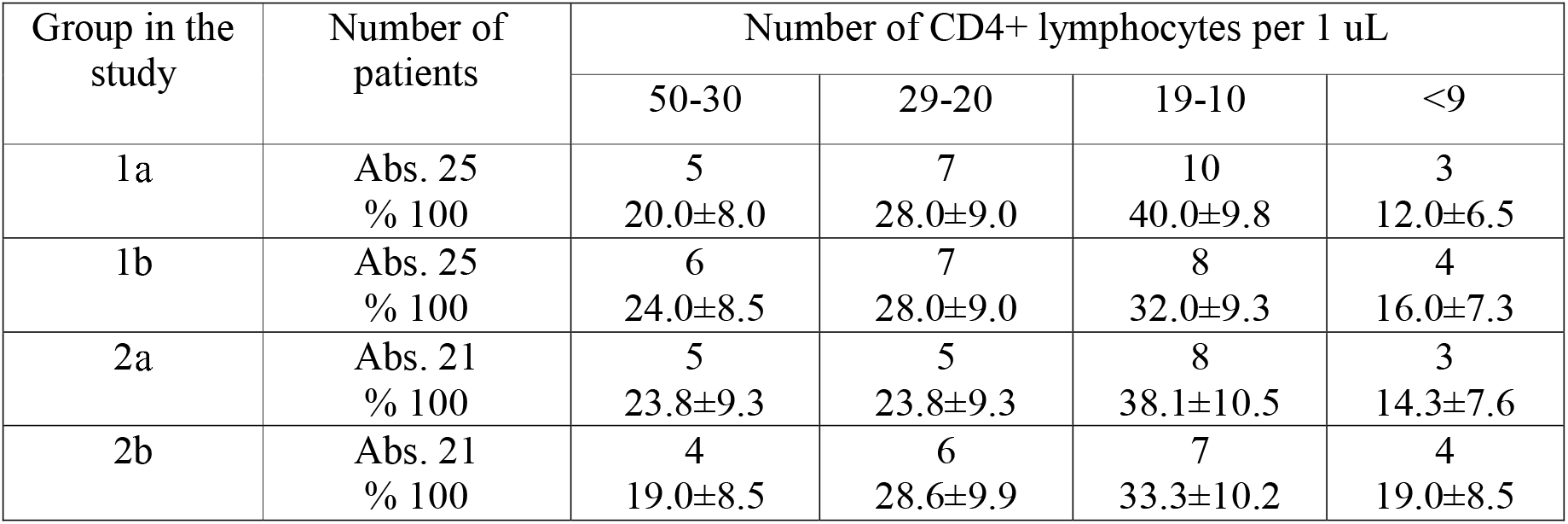
Distribution of patients in groups of the study by the number of CD4+ lymphocytes per 1 uL of blood (M±m)

As shown in Table 1, in groups 1a, 1b, 2a and 2b, the variable of CD4+ lymphocytes number per 1 uL of blood practically did not differ from each other.

In group 1a, in 20.0% of patients, the number of CD4+ lymphocytes was in the range of 50-30 cells /uL of blood, in 28% – 20-29, in 40% – 19-10 and in 12% – less than 9, and in group 1b, respectively: in 24%, 28%, 32% and 16 (p>0.05). In group 2a, the number of CD4+ lymphocytes in 23.8% of patients was in the range of 50-30 cells/uL of blood, in 23.8% – 20-29, in 38.1% – 19-10 and in 14.3% – less than 9, and in group 2b, respectively: in 19.0%, 28.6%, 33.3% and 19.0 (p>0.05).

The mean number of CD4+ lymphocytes per 1 uL of blood was also similar and amounted to 24.7±0.29 cells/uL of blood in group 1a and 29.4±0.44 in group 1b, and in patients of groups 2a and 2b, was respectively: 20.3±0.45 and 28.3±0.33 (p>0.05). At the same time, the viral load in patients of the observed groups was more than 500 000 HIV RNA copies/mL of blood.

Thus, in patients with comorbidity of TRS, HVP and CVP or TRS, CMVP with CVP and without CVP, stage 4 of HIV infection in the phase of progression and absence of ART there is a significant decrease in the number of CD4+ lymphocytes in the blood (from 50 cells and below in 1 uL of blood) with mean number of CD4+ cells not exceeding 30 cells/uL of blood. This indicates a pronounced immunodeficiency, which apparently determines the uniformity of low immune response to pathogens of various etiologies and determines the similarity in the prevalence of pathological lesions and clinic-radiological manifestations of comorbid disease.

In patients of the observed groups, TRS was resulted in generalization of tuberculosis with multiple extrapulmonary specific lesions of various organs, confirmed by isolation of MBT from the diagnostic material of various organs. In patients of group 1a, two organs were affected in 11 patients, three - in 6, four - in 2 and five - in 2, and in group 2a, respectively: two organs - in 10 and three - in 8 and, four - in 1 and five - in 1. The same frequency of extrapulmonary lesions was in groups 1b and 2b where two organs were affected in 12 and 8 patients, three - in 5 and 6, four - in 2 and 5, five - in 2 and 1. The most frequent extrapulmonary localizations in patients of the observed groups were the intestine, mesenteric lymph nodes and peripheral lymph nodes, central nervous system and genitourinary system. Lesions of the spleen, bones and joints were somewhat less common, and lesions of the thyroid gland, adrenal glands, pericardium, inner ear, were found in few patients.

Consequently, in patients with comorbidity of TRS, HVP and CVP or TRS, CMVP with CVP and without CVP, with stage 4 of HIV infection in the phase of progression and absence of ART, the severity of clinical manifestations and course of comorbid disease is largely determined by the generalized nature of tuberculosis against the background of severe immunodeficiency which determines similarity of clinical manifestations of the diseases and hampers their recognition due to the simultaneous combination of several viral pathologies which requires a complex etiological diagnosis of the specific diseases.

The clinical picture of the diseases in patients from the observed groups practically did not differ and was characterized by a pronounced intoxication syndrome and general inflammatory changes with weight loss, adynamia, headache, myalgia, neuropathy, encephalopathy, palpitations, pallor of the skin, fever, chills, signs of inflammation in laboratory tests, decreased saturated oxygen and increasing pulmonary-cardiac insufficiency, that indicates an infectious-toxic shock in the comorbid pathology. All the changes were also combined with symptoms of damage to other organs and systems. All patients of groups 1a and 1b with HVP had typical grouped vesicular eruptions on the mucous membranes (lips, mouth, eyes and genitals) or on various skin areas. In groups 2a and 2b with CMVP, there were rashes on the mucous membranes and on skin represented by petechial or vesicular bullous rash and, in 12 patients, a central nervous system lesion in the form of meningoencephalitis was detected. The clinical picture of inflammatory changes in the respiratory system of the patients also did not differ significantly between the groups and was characterized by shortness of breath, cough, bronchospasm, mucopurulent sputum and presence of different-sized wheezing in the lungs.

Patients of groups 1a and 2a had typical COVID-19 symptoms, anosmia, dysgeusia and sensorineural hearing loss, hypoxemia, the DIC syndrome, thrombosis or thromboembolism, and in some cases - the Guillain-Barre syndrome. However, these clinical manifestations of varying frequencies were also seen in patients without COVID-19, which is largely the result of late stage of HIV infection with severe immunodeficiency and severe course of comorbid disease.

Thus, in patients with comorbidity of TRS, CVP, HVP or TRS, CPMP with CVP and without, stage 4B of HIV infection with immunodeficiency in the phase of progression and absence of ART, the clinical picture, that is characterized by intoxication syndrome, general inflammatory and respiratory manifestations, is almost similar and nonspecific, so it is possible to recognize the clinical course characteristics of the comorbid disease only if the specific pathogen is detected.

On the CT-picture of the chest organs in patients of the observed groups, a complex of simultaneous overlap of four pathological syndromes is visualized: dissemination, pleural pathology, increased pulmonary pattern and adenopathy. Dissemination syndrome, represented by foci of various sizes (from small to large) and intensity (from low to high) with a tendency to merge and form inhomogeneous infiltrates with destruction of lung tissue and bronchogenic contamination. Pleural lesion syndrome was observed and manifested by compaction of the interlobular and parietal pleura in more than half of patients, it coincided with development of exudative pleurisy or pleural empyema. The syndrome of strengthening and deformation of the pulmonary pattern, which had a “mesh” character due to development of interstitial pneumonia and lymphohematogenic dissemination with a diffuse decrease in the lung tissue transparency, development of cystic-dystrophic changes and areas of consolidation of the “frosted glass” type. The adenopathy syndrome is represented by bilateral enlargement of the intra-thoracic lymph nodes with infiltrative changes along the periphery.

In patients of the observed groups, an important role was played by simultaneous overlap of several pathologies and by changes having developed in the lungs during the late stages of HIV infection with immunodeficiency including those directly related to HIV infection itself in the form of lymphoid interstitial pneumonia, nonspecific interstitial pneumonia, primary pulmonary hypertension and high frequent COPD leading to development of emphysema and cystic-dystrophic changes while, at the same time, development of such changes associated with manifestations of VP itself is not excluded [5, 10, 19].

In these cases, the lung lesion area in all patients was 80-100% and was practically comparable between groups. It is not possible to differentiate these changes having been visualized on CT of the chest organs according specific pathologies due to similarity of the CT-pictures, while diagnosis is possible only on the basis of microbiological, virological and molecular genetic determination of the specific pathogens etiology.

As examples, we present CT scans of the chest organs of patients with stage 4B HIV infection with immunodeficiency in the phase of progression and in the absence of ART that illustrate similarity of visualization of the pathological changes in groups 1a and 1b (Fig. 1A and 1B) and in groups 2a and 2b (Fig. 2A and 2B).

**Figure 1.**
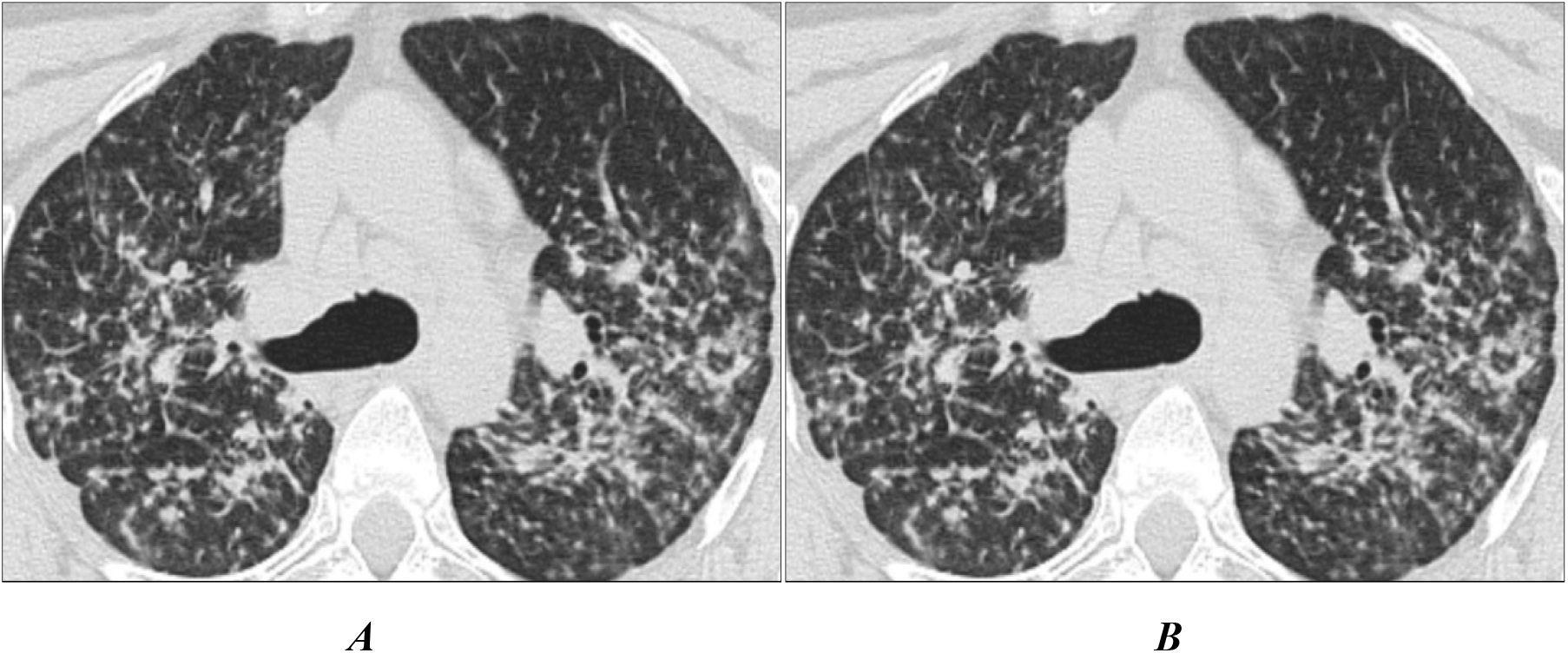
The CT of the chest organs. Axial projection, pulmonary window mode. Pannel A represents a patient with stage 4B of HIV infection with immunodeficiency in the progressive phase without ART and having been verified comorbidity of TRS, HVP and CVP. Pannel B represents a patient with stage 4B of HIV infection with immunodeficiency in the progressive phase without ART and having been verified comorbidity of TRS and HVP.

**Figure 2.**
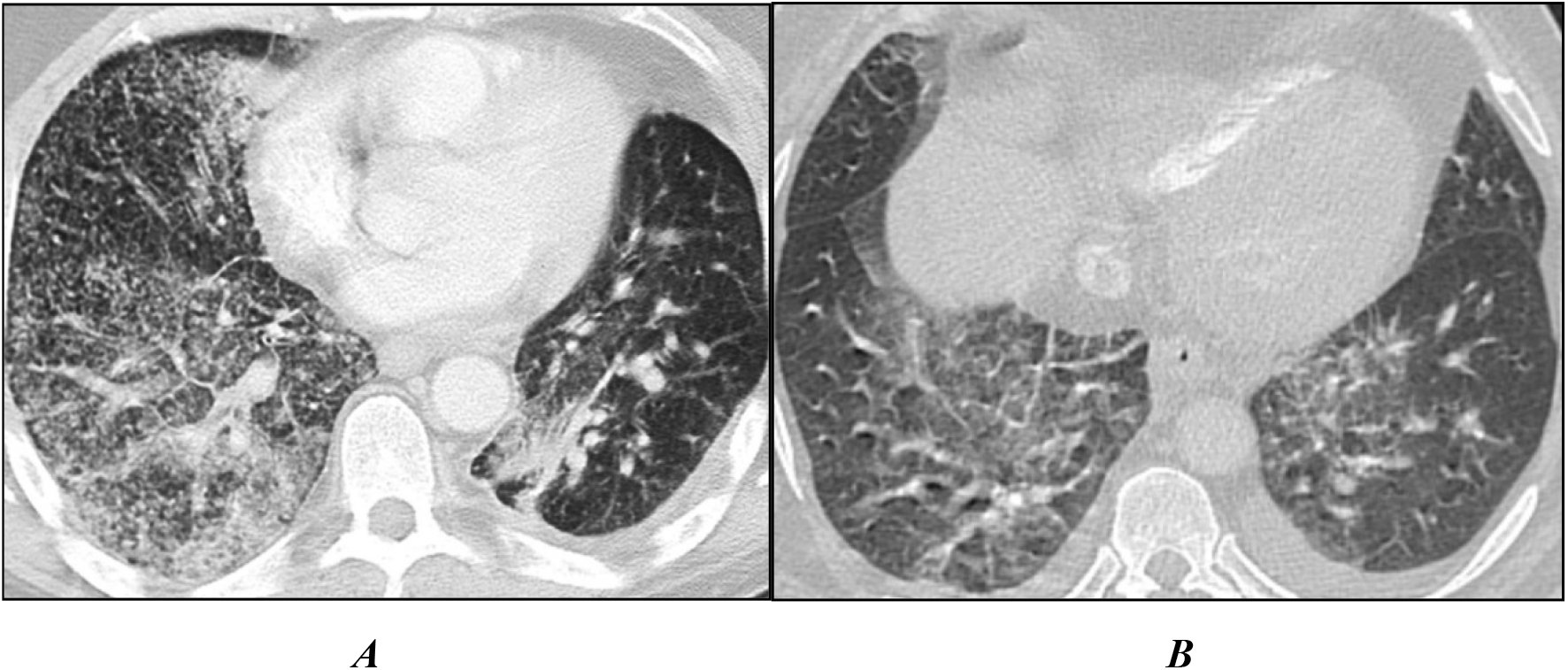
CT of the chest organs. Axial projection, pulmonary window mode. Pannel A represents a patient with stage 4B of HIV infection with immunodeficiency in the progressive phase without ART and having been verified comorbidity of TRS, CMVP and CVP. Pannel B represents a patient with stage 4B of HIV infection with immunodeficiency in the progressive phase without ART and having been verified comorbidity of TRS and CMVP.

As can be seen in Figs. 1 and 2, four syndromes of the same type are visualized on the CT scans of the chest organs: dissemination, pleural pathology, strengthening and deformation of the pulmonary pattern with areas of “frosted glass” and adenopathy.

Thus, the CT scans of the chest organs in patients with comorbidity of TRS, GVP and CVP or TRS, CPMP with CVP and without CVP stage 4 of HIV infection in the phase of progression and in absence of ART with immunodeficiency, the same type of syndromes of several overlapping pathologies are visualized. The pathologies could not be differentially diagnosed and the cases require timely diagnosis using microbiological, molecular genetic and immunological studies to detect *MBT, HVS-1, CMVH and SARS-CoV-2* which is necessary for adequate timely etiological treatment.

## Discussion

Comorbidity of TRS, HVP, CMVP and CVP in patients with late-stage HIV infection in the phase of progression and in absence of ART was diagnosed in patients 5-8 years after the recognition of HIV infection, they were suffering from drug addiction, drunk alcoholic beverages and smoked tobacco products and had concomitant pathology of viral hepatitis B or C and COPD in 66.3% of cases. At the same time, the disease was characterized by severe immunodeficiency and generalization of tuberculosis with multiple extrapulmonary lesions. The latter had determined similarity of clinical manifestations and visualization of CT changes in cases of that comorbidity which hampers their differential diagnostic due to the simultaneous overlap of several pathologies with similar clinical manifestations and requires a complex etiological diagnosis of the specific diseases to prescribe timely comprehensive treatment and to reduce the lethality of this severe contingent of patients. At the same time, it is not possible to differentiate the diseases of the respiratory system according to results of clinical and radiation research methods which requires development of new approaches in the complex diagnosis of comorbidity of TRS and VP of various etiologies applicable in the offices of tuberculosis care for HIV-infected people who must undergo regular examination for timely detection of COVID-19.

## Conclusion

Patients with TRS and HIV infection who are registered in the office of tuberculosis care for HIV-infected in the ATD represent an epidemiological group of high risk from COVID-19 infection and CVP disease and, if severe immunodeficiency, both for HVP and CMVP. They should be regularly subjected to preventive examinations for timely detection of COVID-19 aimed at their emergency isolation and treatment to prevent infection of persons that might be in contact with them.

## Data Availability

All data produced in the present work are contained in the manuscript

## Conflict of Interests

The authors declare that there is no conflict of interest.

## Authors’ contribution

The authors declare the compliance of their authorship according to the international ICMJE criteria. All authors made a substantial contribution to the conception of the work, acquisition, analysis, interpretation of data for the work, drafting and revising the work, final approval of the version to be published and agree to be accountable for all aspects of the work.

## Ethics approval

The study was approved by the local ethics committee of FSBEI HE The A.I. Yevdokimov Moscow State University of Medicine and Dentistry of the Ministry of Health of Russia (protocol №677, 7.10.2022). The approval and procedure for the protocol were obtained in accordance with the principles of the Helsinki Convention

## Notes

### Competing Interest Statement

The authors have declared no competing interest.

### Funding Statement

This study did not receive any funding

### Author Declarations

Ethics committee of the Federal State Budgetary Educational Institution of Higher Education A.I. Evdokimov Moscow State Medical University of the Ministry of Health of the Russian Federation gave ethical approval for this work

## References

1. Cilloniz C, Torres A, Polverino E, Gabarrus A, Amaro R, Moreno E, Villegas S, Ortega M, Mensa J, Marcos M, Moreno A, Miro J. Community-acquired lung respiratory infections in HIV-infected patients: microbial aetiology and outcome. European Respiratory Journal. 2014;43(6):1698–1708. 10.1183/09031936.00155813.

2. Figueiredo-Mello C, Naucler P, Negra M, Levin A. Prospective etiological investigation of community-acquired pulmonary infections in hospitalized people living with HIV. Medicine (Baltimore). 2017;96(4): pp. 57–78. 10.1097/MD.0000000000005778

3. Belyakov NA, Rassokhin VV. HIV infection and comorbid conditions: monograph. St. Petersburg: Baltic Medical Education Center, 2020, 680 p. (in Russian) https://expose.gpntbsib.ru/expose/vnp-40ada057/book/D2020-2490878857701

4. Fazylov VH, Manapova ER, Akifyev VO. Clinical and epidemiological characteristics of secondary diseases in HIV-infected patients in the conditions of inpatient care. Infectious diseases: news, opinions, training. 2020;9(4):81–87 (In Russian) 10.33029/2305-3496-2020-9-4-81-87

5. Guidelines for Prevention and Treatment of Opportunistic Infections in HIV-Infected Adults and Adolescents with HIV. Recommendations from the Centers for Disease Control and Prevention, the National Institutes of Health, and the HIV Medicine Association of the Infectious Diseases Society of America, 2021, 479 p. https://clinicalinfo.hiv.gov/sites/default/files/guidelines/documents/Adutl_Ol.pdf

6. Mishina AV, Mishin VYu, Ergeshov AE, Sobkin AL, Romanov VV, Kononec AS. Features of clinical manifestations and diagnostics of a combination of respiratory tuberculosis with opportunistic lung infections in adult patients with late stages of HIV infection with immunodeficiency. Consilium Medicum. 2020;22(11):78–86 (in Russian) 10.26442/20751753.2020.11.200184

7. Azovtseva O.V. Features of the course of herpetic infection on the background of HIV infection. HIV infection and immunosuppression. 2010;2(3):37–41 (In Russian).

8. Sklyar LF, Markelova EV, Borovskaya NA, Zima LG, Gaponenko EK. Clinical and immunological features of herpesviral diseases in case of HIV-infection. Pacific Journal. 2010;(3):62–64 (In Russian).

9. Anaedobe CG, Ajani TA. Co-infection of Herpes Simplex Virus Type 2 and HIV Infections among Pregnant Women in Ibadan, Nigeria. J Glob Infect Dis. 2019 Jan-Mar;11(1):19–24. 10.4103/jgid.jgid-56-18

10. Bartlett G., Redfield R., Pham P. Bartlett’s Medical Management of HIV Infection. Oxford Univercity Press. 2019, 864 p. 10.1093/med/9780190924775.001.0001

11. Shakhgildyan VI, Yandrinskaya MS, Orlovsky AA, Shipulina OY, Domonova EA, Tishkevich OA, Ermak TN, Yrovaya EB. The concentration of CMV DNA in biological materials is the key to the diagnosis of cytomegalovirus pneumonia in patients with HIV infection. Journal of Infectology (S1). 2019;11(3):109–117 (In Russian) 10.22625/2072-6732-2019-11-3S1.

12. Shakhgildyan VI, Yadrikhinskaya MS, Orlovsky AA, Domonova EA, Shipulina OY, Tishkevich O., Ermak TN, Yrovaya EB. Virial, but not-covid pneumonia in patients with HIV infection. Epidemiology and infectious diseases. 2021:11(3):69–77 (In Russian) 10.18565/epidem.2021.11.3.69-77

13. Gervasoni C, Meraviglia P, Riva A, Giacomelli A, Oreni L, Minisci D, Atzori C, Ridolfo A, Cattaneo D. Clinical features and outcomes of HIV patients with coronavirus disease 2019. Clin Infect Dis. 2020 May 14, pii: ciaa 579. 10.1093.cid.ciaa579

14. Hopkins J. COVID-19 in patients with HIV: clinical case series. Lancet HIV. 2020 Apr 15;1–9. 10.1016/

15. Hoffmann C, Casado J, Härter G, Vizcarra P, Moreno A, Cattaneo D, Meraviglia P, Christoph D., Schabaz F., Grunwald S, Gervasoni C. Immune deficiency is a risk factor for severe COVID-19 in people living with HIV. HIV Medicine. 2021 May;22(5);372–378. 10.1111/hiv.13037

16. Gauss AA, Klimova NV. Rentrenomorphological features of the COVID-19 and HIV infections. HIV infection and immunosuppression. 2021;13 (2):77–84 (In Russian) 10.22328/2077-9828-2021-13-2-77-84

17. Kravchenko A.V., Kuimova U.A., Kanestri V.G., Goliusova M.D., Kulabukhova E.I. Clinical course and approaches to therapy of patients with combined infection (HIV infection and COVID-19). Epidemiology and infectious diseases. 2021;4;20–24 (In Russian). 10.18565/epidem.2021.11.4.20-4

18. Federal clinical recommendations on diagnosis and management of TB in HIV-infected patients. Moscow-Tver, Triada, 2014, 56 p. (In Russian) https://aidsomsk.ru/sites/default/uploads/_-.pdf

19. Clinical recommendations. HIV infection in adults. National Association of Specialists in the Prevention, Diagnosis and Treatment of HIV Infection. 2020, 230 p. (In Russian). https://rushiv.ru/klinicheskie-rekomendatsii-vich-infektsiya-u-vzroslyh-2020/

20. Temporary methodical recommendations of the Ministry of Health of the Russian Federation. Prevention, diagnosis, and management of the novel coronavirus infection (COVID-19). Version 16 (18.08.2022): 248 (in Russian). https://static-0.minzdrav.gov.ru/system/attachments/attaches/000/060/193/original/BMP_COVID-19_V16.pdf

